# Early Outcomes of Preoperative 5-fraction Radiation Therapy for Soft Tissue Sarcoma Followed by Immediate Surgical Resection

**DOI:** 10.1101/2020.03.20.20038885

**Authors:** Shireen Parsai, Joshua Lawrenz, Scott Kilpatrick, Brian Rubin, Cory Hymes, Michele Gray, Nathan Mesko, Chirag Shah, Lukas Nystrom, Jacob G Scott

## Abstract

**Purpose/Objectives:** There are limited data regarding the use of hypofractionated radiation therapy (RT) for soft tissue sarcoma. We report early oncologic outcomes and wound complications of patients undergoing preoperative hypofractionated (5 fraction) RT followed by immediate surgical resection.

**Materials/Methods:** An IRB-approved database of patients treated with preoperative RT for soft tissue sarcoma was queried. Patients treated with a hypofractionated dosing regimen followed by immediate (within 7 days) planned wide surgical resection were identified.

**Results:** Between 2016 to 2019, sixteen patients met eligibility criteria. The median clinical follow-up was 10.7 months (range 1.7-33.2). The median patient age was 64 years old (range 33-88). Ten of the sarcomas were located in the lower extremity, 4 in the upper extremity, and two were located in the trunk. Five patients had metastatic disease at diagnosis. The majority of the patients received a total radiation dose of 30 Gy in 5 fractions (range 27.5-40 Gy) on consecutive days. All patients were planned with IMRT/VMAT. The median time to surgical resection following the completion of RT was 1 day (range 0-7 days). The median time from initial biopsy results to completion of primary oncologic therapy was 20 days (range 16-35). Ten patients achieved R0 resection, whereas the remaining 6 patients achieved R1 resection. Of the 13 patients assessed for local control, no patients developed local failure. Five patients developed wound healing complications (31%), of which only three patients (19%) required return to the operating room.

**Conclusions:** Treatment of soft tissue sarcoma with preoperative hypofractionated RT followed by immediate resection resulted in a median of 20 days from biopsy results to completion of oncologic therapy. Early outcomes demonstrate favorable wound healing. Further prospective data with long-term follow-up is required to determine the oncologic outcomes and toxicity of hypofractionated preoperative RT.

## Introduction

Soft tissue sarcomas are rare malignant tumors of mesenchymal origin. The mainstay of treatment is complete surgical resection and radiation therapy (RT), delivered either in the neoadjuvant or adjuvant setting. Local control and survival outcomes are similar when comparing neoadjuvant and adjuvant RT, though the toxicity profiles differ.^1,2^ Pre-operative radiotherapy is known to have a higher rate of wound healing complications, but improved extremity function due to less fibrosis of the periarticular soft tissues.^1^ Conventional neoadjuvant RT regimens consist of 50 Gy in 25 fractions (fx) delivered over five weeks. When including a 3-6 week post-treatment period, time to surgical resection, and therefore completion of primary oncologic therapy, ranges between 8-11 weeks from the time RT is initiated.^1^ Hypofractionated RT, defined as >2.2 Gy delivered per fraction, has been shown to be efficacious in other cancer types, including breast^3–5^, rectal^6^ and prostate cancer.^7^ Prior experiences of preoperative hypofractionated regimens have used 3.5 − 8 Gy per fraction^8–12^. Hypofractionated regimens considerably shorten the entire treatment package time. This allows for quicker time receiving adjuvant therapies, undergoing physical therapy, and re-introduction of the patient back into society with less time off work. In 2014, Kosela-Paterczyk et al. reported on a prospective single-arm study of 272 patients treated with 25 Gray in 5 fractions followed by immediate (within 3-7 days) surgical resection.^12^ Local control rate was lower than expected at 81% with a median follow-up of 35 months. However, the wound complication rate of 7% was deemed favorable. Kalbasi et al reported on a single arm prospective study of patient treated with 30 Gy in 5 fractions, followed by delayed surgery (within 2-6 weeks).^13^ The local control rates reported on 35 evaluable patients was 94.3% at 2 years. In this study, major wound complications occurred in 32%. At our institution, we have selectively used a high-dose preoperative hypofractionated regimen for patients with soft tissue sarcomas outside the abdomen and pelvis followed by immediate surgery. This represents a further advance combining the novelties of the regimens previously published by Kosela-Paterczyk et al and Kalbasi et al..^12, 13^ Given the paucity of data, we aimed to assess our early clinical, toxicity and histologic data of patients receiving hypofractionated RT followed by immediate surgical resection.

## Materials and Methods

An institutional review board-approved database of patients treated with preoperative 5-fraction radiation regimens between 2016-2019 was queried. Patients with soft tissue sarcomas of the extremities or trunk treated with preoperative hypofractionated radiation therapy were included, whereas retroperitoneal sarcomas were excluded. The decision to use hypofractionated dosing regimen with immediate surgery was made as part of a multidisciplinary discussion as well as sarcoma tumor board. All patients were treated at a single institution. Following surgical resection, patients were seen for surveillance every 3-4 months for the first three years. Surveillance imaging included a CT chest without contrast, as well as imaging of the primary tumor with MRI.^14^

### Radiation Target Delineation and Treatment Planning

All patients received hypofractionated preoperative radiation therapy over 5 fractions. Tumor volumes were generally designed as was done on RTOG 0630.^15^ The planning CT was registered with preoperative MRI imaging performed before or at the time of simulation. The gross tumor volume (GTV) was defined based on preoperative MRI with sequence based on histology. The CTV was drawn to respect anatomical barriers to tumor spread. The PTV was generated by adding 5 mm to the CTV. Daily image guidance with cone beam computed tomography was employed. Both step and shoot IMRT and VMAT radiation techniques were allowed. We attempted to limit bone in proximity to PTV to V15 Gy < 50% and to spare a strip of skin > 2 cm to less than 10 Gy. We also consulted with our orthopaedic surgeons to avoid hotspots in the skin where the surgical wound was planned.

### Surgery and Reconstruction

Surgical resection was performed by an orthopaedic oncologic surgeon (authors blinded for review), 0 − 7 days following completion of the preoperative RT. Timing decisions were based upon patient preference and surgeon schedules. Wound healing complications were defined as previously described by O’Sullivan, et al..^1^ Major wound complications were those requiring return to the operating room, admission for IV antibiotics and wound packing > 120 days, or aspiration of seroma.

### Statistical Analysis and Definitions

Descriptive statistics were used to describe local failure, survival, toxicities, and wound complications of treatment. Local failure was assessed radiographically on surveillance MRI. Local failure was reported for patients that had > 3 months of radiographic follow-up.

## Results

### Patient, Tumor, and Treatment Characteristics

A total of sixteen patients met the study eligibility criteria. A summary of the patient, tumor, and treatment characteristics is included in Table 1. Ten patients had diagnoses of soft tissue sarcomas of the upper extremity, 4 lower extremity, and the remaining two patients had trunk sarcomas. The majority of the patients had localized disease, however four patients had metastatic disease at diagnosis. Of the four patients who had metastatic disease at diagnosis, three patients had undergone prior chemotherapy before preoperative radiotherapy and surgical resection. Most patients received 30 Gy in 5 fractions, with a dose range of 27.5 Gy-40 Gy. The median time from completion of radiation to surgical resection was 1 day (range 0-7). Median time from initial biopsy results to completion of primary oncologic therapy was 20 days (range 16-35) as shown in Figure 1. The median time from initial biopsy to completion of primary oncologic therapy was 27 days (range 21-38).

**Table 1.** Patient, tumor, and treatment characteristics.

|  |  |
| --- | --- |
| **Age** | Median 64 years (range 33-88) |
| **Gender** |  |
| Male | 9 |
| Female | 7 |
| **Site** |  |
| Extremity | 14 |
| Upper Extremity | 4 |
| Lower Extremity | 10 |
| Trunk | 2 |
| **Histology** |  |
| Dedifferentiated liposarcoma | 1 |
| CIC-rearranged sarcoma | 1 |
| Extraskeletal myxoid chondrosarcoma | 1 |
| Myxofibrosarcoma | 4 |
| Mxoid Liposarcoma | 3 |
| Pleomorphic leiomyosarcoma | 1 |
| Synovial Sarcoma | 2 |
| Undifferentiated Pleomorphic/ spindle cell sarcoma | 3 |
| **Grade** |  |
| Grade 2 | 8 |
| Grade 3 | 3 |
| **Greatest dimension on final pathology** |  |
| ≤ 5cm | 3 |
| > 5cm and ≤ 10cm | 9 |
| > 10cm and ≤ 15cm | 3 |
| > 15cm | 1 |
| **Metastatic at diagnosis** | 4 |
| **PTV size** | Median 485cm<sup>2</sup> (range 56-4353) |
| **Radiation Technique** |  |
| Step and Shoot | 3 |
| VMAT | 13 |
| **Radiation Dose** |  |
| 27.5Gy 5 fx | 1 33-88) |
| 30Gy 5 fx | 14 |
| 40Gy 5 fx | 1 |
| **Prior unplanned excision** | 4 |
| **Wound closure** |  |
| Primary | 13 |
| Regional/Pedicled flap** | 2 |
| Free flap | 1 |
| Skin graft** | 1 |
| **Pathologic margin status\*\*\*** |  |
| R0 | 10 |
| R1 | 6 |

\* Grade not available for 5 patients;
\*One patient underwent closure with regional pedicle and skin graft;
\*\*R0: gross total resection; R1: microscopically positive margins

**Table 2.**
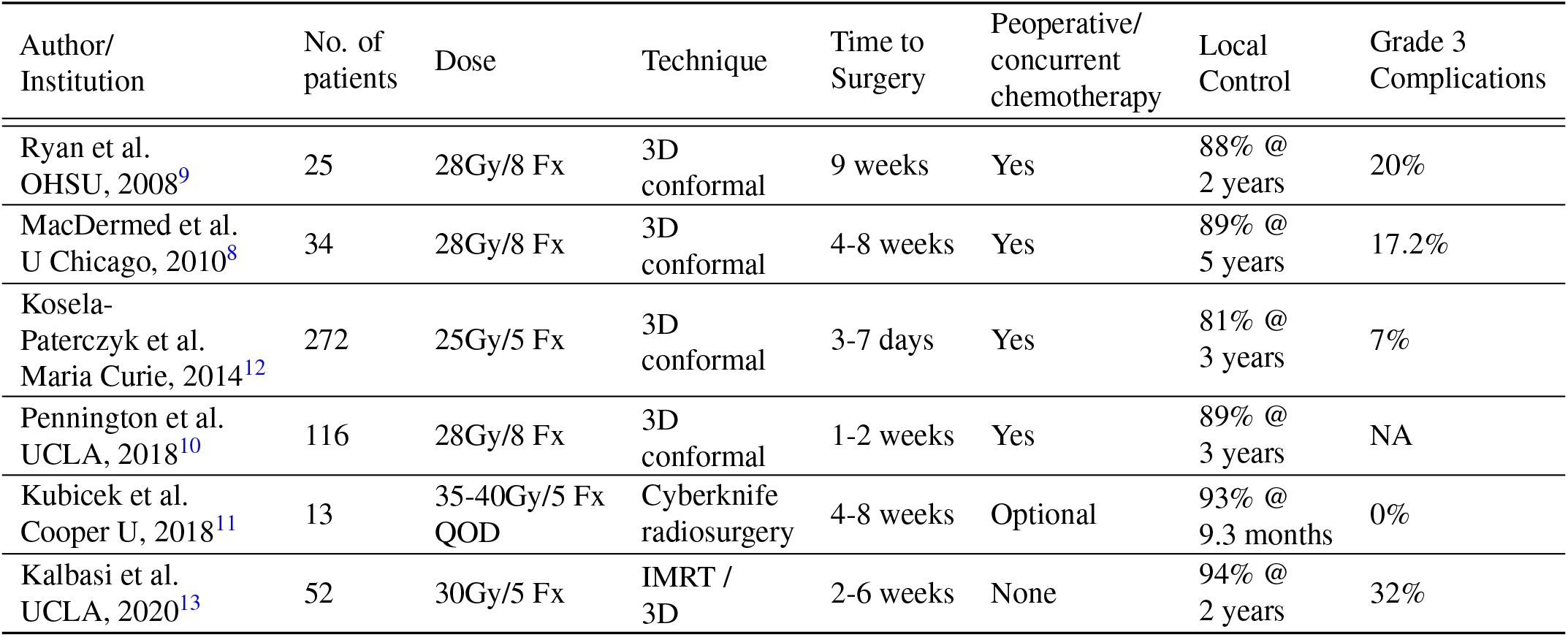
Summary of other studies employing hypofractionated preoperative radiotherapy for soft tissue sarcoma.

| Author/<br>Institution | No. of<br>patients | Dose | Technique | Time to<br>Surgery | Preoperative/<br>concurrent<br>chemotherapy | Local<br>Control | Grade 3<br>Complications |
| --- | --- | --- | --- | --- | --- | --- | --- |
| Ryan et al.<br>OHSU, 2008<sup>9</sup> | 25 | 28Gy/8 Fx | 3D<br>conformal | 9 weeks | Yes | 88% @<br>2 years | 20% |
| MacDermid et al.<br>U Chicago, 2010<sup>8</sup> | 34 | 28Gy/8 Fx | 3D<br>conformal | 4-8 weeks | Yes | 89% @<br>5 years | 17.2% |
| Kosela-Paterczyk et al.<br>Maria Curie, 2014<sup>12</sup> | 272 | 25Gy/5 Fx | 3D<br>conformal | 3-7 days | Yes | 81% @<br>3 years | 7% |
| Pennington et al.<br>UCLA, 2018<sup>10</sup> | 116 | 28Gy/8 Fx | 3D<br>conformal | 1-2 weeks | Yes | 89% @<br>3 years | NA |
| Kubicek et al.<br>Cooper U, 2018<sup>11</sup> | 13 | 35-40Gy/5 Fx<br>QOD | Cyberknife<br>radiosurgery | 4-8 weeks | Optional | 93% @<br>9.3 months | 0% |
| Kalbasi et al.<br>UCLA, 2020<sup>13</sup> | 52 | 30Gy/5 Fx | IMRT /<br>3D | 2-6 weeks | None | 94% @<br>2 years | 32% |

**Figure 1.**
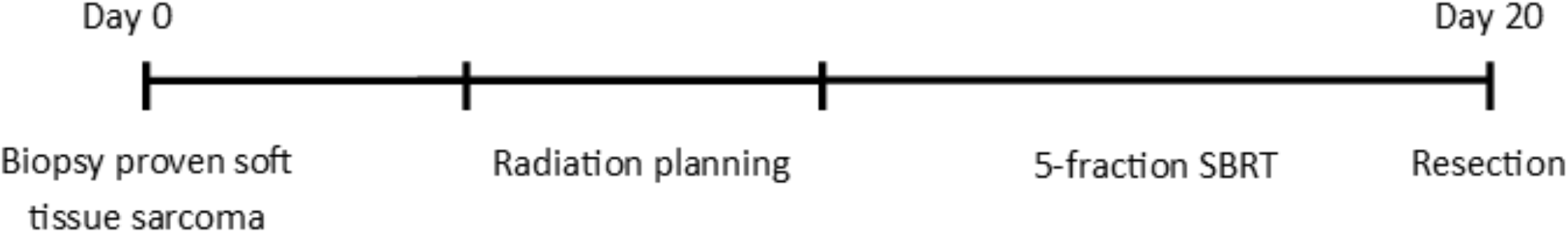
Treatment Schema for preoperative hypofractionated radiotherapy followed by immediate surgical resection. The median time from initial biopsy results to surgical resection was 20 days. Surgical resection occurred within 7 days of completion of radiation therapy.

### Oncologic Outcomes

The median follow-up time was 10.7 months (range 1.7 − 32.0). Within the follow-up time, 2/16 patients died. Both of these patients had metastatic disease at the time of radiation and surgical resection, and continued to have distant progression, ultimately leading to demise. Within the follow-up time, no local failures were observed in 13 evaluable patients. Gross total resections (R0) were achieved in 10 patients. In the remaining six patients, microscopically positive margins were identified. Five patients had planned marginal (R1) resections. One patient with myxofibrosarcoma had persistently positive margins outside of the radiation field following re-resection. This patient underwent postoperative re-irradiation conventional fractionation to 66 Gy in 33 fractions.

### Wound Complications and Toxicity

No acute or late grade 3 or higher radiation-related toxicities were observed, including lymphedema, fibrosis or joint stiffness. Major wound complications requiring re-operation occurred in 3/16 patients (18.8%). Minor wound complications occurred in 2/16 patients (12.5%). Major and minor wound complications included dehiscence, skin necrosis, and seroma. Figure 2 demonstrates representative images of wound complications.

**Figure 2.**
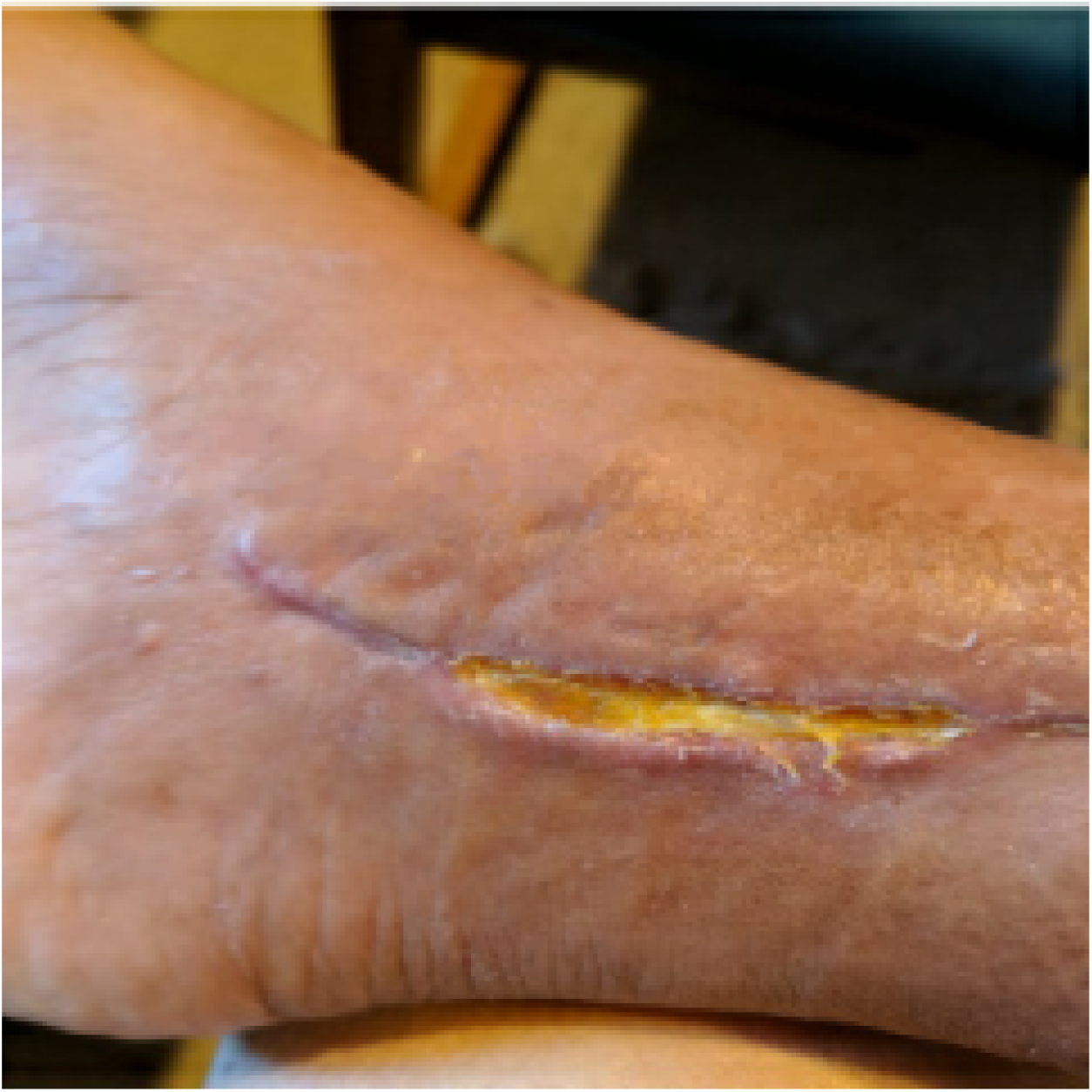
Minor wound complication. This is a patient with skin necrosis which ultimately led to cellulitis requiring short-term IV antibiotics, and was characterized as a minor complication. This patient did not require reoperation.

## Discussion

We present early oncologic and toxicity outcomes of treating soft tissue sarcomas with a 5-fraction preoperative hypofractionated radiotherapy courses followed by immediate surgical resection. This strategy significantly reduces the overall treatment time as compared to conventional preoperative radiation therapy followed by delayed surgery, which generally ranges between 8-12 weeks. Our data demonstrate the median time from obtaining the biopsy results to surgical resection of the primary tumor, including preoperative radiation, was less than three weeks. In this way, the treatment package time was reduced by 5-9 weeks. Theoretically, decreased treatment completion time allows less time for the primary tumor to develop micrometastatic disease. Additionally, for metastatic patients shortening the treatment course allows quicker resumption of systemic therapy. Patients will also have reduced less absences from work and quicker resumption of activities of daily living.

With only 3 of 16 patients requiring re-operation for wound healing complications, our data demonstrate a rate comparable to that reported in the literature for standard fractionation radiotherapy.^16^ On the NCIC study reported by O’Sullivan et al, acute wound complications were recorded in 35% of patients who received preoperative radiotherapy vs 17% in those that received postoperative radiotherapy.^17^ Of the patients who underwent preoperative radiotherapy, 16% of patients required reoperation due to treatment complications.^1^ This is comparable to our series, in which nearly 19% of patients required a reoperation. To date, most preoperative short course radiotherapy series have utilized 3D conformal techniques over more modern techniques. Older 3D conformal techniques, were employed in the original NCIC study of preoperative vs postoperative therapy, demanded larger target volumes without dose painting to avoid normal structures. In our series, we uniquely employed either step and shoot IMRT (intensity modulated radiation therapy) or VMAT (volumetric arc therapy) with image guidance for all patients. O’Sullivan et al demonstrated the use of IMRT with image guidance and that restricting radiation dose to future surgical skin flaps and bone reduced the risk of toxicity.^17^ Wang et al (RTOG 0630) further used image guidance to reduce target volumes, which led to decreased late toxicities without an increase in the risk of recurrence.^15^ The radiation techniques employed in this study were modeled after RTOG 0630. Surgical efforts to decrease wound healing complications in this study included immediate surgical resection, layered closure when possible, use of deep drains to prevent seroma formation, and standard use of incisional wound vacuums.

Local control in this study is favorable, although our follow-up interval is too short to make definitive statements. No local failures observed within the follow-up period. Maturation of data is important to ensure longer term control remains favorable. A recent study of hypofractionated preoperative radiation (30 Gy in 5 fractions) reported by Kalbasi et al demonstrated a 2-year local control rate of 94.3%.^13^ This can be compared to the historical standard 5-year local control of 93%.^1^ Table **??** summarizes the local control rates for prior experiences with hypofractionated preoperative radiotherapy. The largest reported series to date demonstrated an unexpectedly low 3-year local control rate of 81%.^12^ This may be attributed to a low total radiation dose of 25 Gy in 5 fractions without chemotherapy. We chose to treat all patients with doses greater than 25 Gy (EQD2 = 40, calculated at alpha-beta ratio of 3) due to the lower local control observed in that report, though in the future, we imagine that patients will be given personalized fractionation based on their tumor genomics.^18^ Most patients received radiation doses of 30 Gy in 5 fractions (EQD2= 54, calculated at alpha-beta ratio of 3) prior to surgery with favorable toxicity profiles. In our series, immediate surgical resection was employed in an effort to shorten the overall treatment course, similar to the treatment regimen published by Kosela-Paterczyk et al.^12^

As a retrospective study design, this investigation has inherent limitations. The primary limitation is a selection bias of patients who have tumors felt to be amenable to hypofractionated radiation therapy and excluded cases at high-risk for toxicity (ex. circumferential tumor close to bone). Additional limitations include a small number of patients as well a heterogeneous group in regards to stage, histology, location and range of RT doses. Further prospective studies are required employing modern techniques of preoperative hypofractionated radiotherapy and immediate surgical resection.

## Conclusion

Our use of hypofractionated preoperative RT for soft tissue sarcoma with immediate resection resulted in a median of 20 days from biopsy results to resection of the primary tumor, reducing the time from diagnosis to completion of primary oncologic therapy by nearly 3 months for these patients. Early outcomes demonstrate an acceptable rate of wound healing complications comparable to standard therapy. Further prospective data with long-term follow-up is required to determine the oncologic outcomes and toxicity of preoperative hypofractionated RT.

## Data Availability

Institutional data is available as needed.

